# Disease-Specific Trends and Ethnic Inequities in In-Hospital Cardiovascular Mortality in Suriname, 2013–2022

**DOI:** 10.64898/2026.11.08.26367569

**Authors:** Ritesh Dhanpat, Rakesh Gajadhar Sukul, Angele Mendeszoon, Vanita Jairam, Jerry Toelsie, Robbert Bipat

**Affiliations:** Faculty of Medical Sciences, Anton de Kom University of Suriname, Paramaribo, Suriname; Ministry of Health, Wellness and Labor, Paramaribo, Suriname; ’s Lands Hospital, Paramaribo, Suriname; Academic Hospital Paramaribo, Paramaribo, Suriname

**Keywords:** cardiovascular disease, cerebrovascular disease, ischemic heart disease, heart failure, in-hospital mortality, health equity, ethnic disparities, Suriname

## Abstract

**Objective:** To examine temporal trends in in-hospital mortality among patients hospitalized with heart failure, ischemic heart disease, and cerebrovascular disease in Suriname from 2013 to 2022, and to assess differences according to age, sex, and ethnicity.

**Methods:** Conducted a retrospective hospital-based observational study using routinely collected discharge data from three major public hospitals in Suriname. These hospitals accounted for approximately 71.8% of national hospital admissions. Admissions with a primary diagnosis of HF (ICD-10 I50), IHD (I20–I25), or Stroke (I60–I69) were included. The primary outcome was in-hospital mortality. Separate multivariable logistic regression models were fitted for each disease, adjusting for calendar year, age, sex, and ethnicity.

**Results:** Among 84,691 admissions, 42,301 were IHD, 24,584 for Stroke, and 17,806 for HF. In-hospital mortality declined significantly across all three conditions, with adjusted odds decreasing by 14% annually for HF (aOR 0.858; 95% CI 0.841–0.875), 6% for IHD (aOR 0.941; 95% CI 0.914–0.969), and 5% for Stroke (aOR 0.947; 95% CI 0.931–0.963). Increasing age was associated with higher mortality in IHD and Stroke. Female sex was associated with lower mortality in HF (aOR 0.835; 95% CI 0.744–0.937). Ethnic disparities were most pronounced for Stroke, with higher mortality among Maroon, Creole, Javanese, and Other groups than Hindustani patients.

**Conclusions:** In-hospital mortality declined across cardiovascular conditions, but improvements were heterogeneous. Pronounced ethnic inequalities in Stroke mortality and age-related gradients highlight the need for equity-focused cardiovascular surveillance and provide a baseline for evaluating future health-system and prevention initiatives in Suriname.

## Introduction

Cardiovascular diseases remain the leading cause of death globally, accounting for approximately 17.9 million deaths annually. Although age-standardized cardiovascular mortality has declined in many high-income countries, the absolute burden continues to increase in low- and middle-income countries (LMICs), driven by population growth, aging, and persistent exposure to preventable risk factors (1–3)

### The Caribbean Transition

The Caribbean region faces a distinct epidemiologic profile. Rapid urbanization, dietary shifts toward processed foods, declining physical activity, and rising obesity prevalence have driven accelerating non-communicable disease burden (4–7). Unlike high-income countries with established cardiovascular prevention systems, many Caribbean nations are navigating the early phase of cardiovascular transition—where disease incidence is rising faster than healthcare system capacity can accommodate(8). Additionally, the Caribbean’s multiethnic populations (South Asian, African, indigenous, and mixed ancestry) have historically exhibited differential cardiovascular risk, yet longitudinal data on whether health systems are reducing this inequity remain scarce(9).

### Policy Context: The HEARTS Initiative as a Timely Response

Suriname exemplifies Caribbean health system challenges. Primary healthcare is delivered through fragmented public (Regional Health Services, Medical Mission) and private sectors, with no unified electronic health record, variable resource allocation, and limited real-time surveillance of acute cardiovascular outcomes (10). Like many Caribbean nations, Suriname faces chronic shortages in cardiovascular specialists, inconsistent access to evidence-based medications, and competing priorities across a resource-constrained system(7).

In December 2021, Suriname’s Ministry of Health officially adopted the HEARTS in the Americas Initiative—a WHO-endorsed primary healthcare strengthening program targeting hypertension detection and control(11). HEARTS focuses on six core components: healthy-lifestyle counseling, evidence-based treatment protocols, access to essential medicines, risk-based management, team-based care, and systems monitoring and evaluation(12,13). Operational implementation began in 2023, with the goal of achieving population-level blood pressure control rates ≥50% by 2025 (14) . However, the baseline health system performance on acute cardiovascular mortality— the ultimate indicator of prevention effectiveness—had not been systematically evaluated. Baseline data are essential for (1) establishing targets against which to measure HEARTS impact,(2) identifying which population groups are at highest risk and require targeted interventions, (3) determining whether HEARTS should be coupled with acute care strengthening strategies, and (4) planning cardiovascular service capacity over the next decade.

This study therefore evaluates health system performance on cardiovascular mortality in the immediate pre-implementation period (2013–2022), providing a baseline against which to measure HEARTS impact and identifying equity gaps that prevention alone may not resolve.

## Methods

### Study design and setting

We conducted a retrospective hospital-based observational cohort study using routinely collected hospital discharge data from three major public hospitals in Suriname: Academic Hospital Paramaribo, ‘s Lands Hospital, and Mungra Medical Center. The study period extended from January 1, 2013, through December 31, 2022. Together, the participating hospitals accounted for approximately 71.8% of national hospital admissions and provide secondary and tertiary care to populations from different geographic areas of Suriname. The study was based on routinely collected administrative hospital data. The unit of analysis was the hospital admission rather than the individual patient.

### Study population

All hospital admissions with a primary discharge diagnosis of Hearts Failure (HF), Ischemic Hearts Disease (IHD), or Stroke during the study period were eligible for inclusion. HF was identified using ICD-10 code I50, IHD using codes I20–I25, and Stroke using codes I60–I69. The diagnostic categories were analyzed separately because they represent clinically heterogeneous conditions with different patterns of presentation and prognosis.

### Diagnostic classification

IHD (I20–I25) included ischemic heart disease diagnoses recorded in the hospital discharge data, including angina, acute myocardial infarction, and other ischemic heart disease diagnoses. Stroke (I60–I69) included cerebrovascular diagnoses recorded in the discharge data, including hemorrhagic and ischemic cerebrovascular conditions, transient ischemic attack, and late effects of cerebrovascular disease. HF (I50) included heart failure diagnoses recorded in the discharge data, including acute and chronic heart failure presentations. The administrative nature of the dataset means that the clinical specificity of individual diagnoses was dependent on the quality and completeness of ICD-10 coding.

### Outcome

The primary outcome was in-hospital mortality, defined as death occurring during the index hospitalization. For descriptive analyses, the crude case-fatality proportion was calculated as the number of admissions resulting in death divided by the total number of admissions within each diagnostic and demographic category. In-hospital mortality reflects short-term mortality among hospitalized patients and should not be interpreted as population-level mortality, disease-specific incidence, or long-term survival.

### Demographic variables

The principal demographic variables were age, sex, and ethnicity. Age was analyzed as a continuous variable in years. Sex was categorized as male or female according to the hospital administrative record. Ethnicity was categorized into five population groups routinely recorded in hospital administrative data: Hindustani, Maroon, Creole, Javanese, and Other. Hindustani patients were used as the reference category in regression models. Ethnicity was treated as a social and population characteristic rather than a biological determinant. The categories may reflect differences in social, cultural, geographic, and structural circumstances. Because ethnicity was obtained from routinely collected administrative records, misclassification or inconsistencies in classification cannot be excluded.

### Length of hospital stay

Length of hospital stay (LOS) was measured in days. Because LOS is determined after admission and may be influenced by disease severity, treatment, discharge practices, and death during hospitalization, it was not included as a primary explanatory variable in the main mortality models. Its association with mortality was examined separately. This distinction is important because early death may itself result in a shorter observed hospital stay, creating potential reverse causation and survivor bias.

### Statistical analysis

Descriptive statistics were used to summarize admissions according to diagnosis, sex, and ethnicity. Crude case-fatality proportions were calculated overall and by ethnic group. Temporal trends in in-hospital mortality were examined using separate multivariable logistic regression models for HF, IHD, and Stroke. In each model, in-hospital mortality was the binary outcome. Calendar year was modeled continuously. The resulting adjusted odds ratio represents the change in the odds of in-hospital mortality associated with each additional calendar year. An aOR below 1.00 therefore indicates declining odds of mortality over time; the approximate annual percentage decline in odds was calculated as (1 ™ aOR) × 100.

Age, sex, and ethnicity were included as covariates. Male sex and Hindustani ethnicity were used as reference categories. Interaction terms between calendar year and demographic variables were evaluated to assess whether temporal mortality trends differed across demographic groups. Interaction terms were retained when statistically significant and when they contributed meaningfully to model fit, according to the prespecified criteria in the analytical framework.

Model fit was assessed using McFadden and Nagelkerke pseudo-R^2^ statistics. The generally low explanatory power of the models was considered when interpreting the findings because the administrative dataset did not contain several clinically important determinants of mortality, including disease severity, comorbidities, vital signs, laboratory findings, imaging findings, treatment, and time to presentation.

All analyses were conducted using IBM SPSS Statistics and JASP. Statistical tests were two-sided, with p<0.05 considered statistically significant. Gemini AI was used to create figures.

### Ethical considerations

The study was conducted in accordance with applicable national regulations and institutional requirements governing the use of routinely collected health data. Ethical oversight was provided by the relevant national review body under approval letter CMWO-14, dated March 25, 2026. The study used retrospectively collected, anonymized hospital data, and the requirement for informed consent was waived in accordance with applicable national legislation and institutional policy.

## Results

### Study population and baseline characteristics

Between 2013 and 2022, 84,691 hospital admissions for Stroke, IHD, and HF were included in the analysis (Table 1). IHD accounted for half of all admissions (50.0%), followed by Stroke (29.0%) and HF (21.0%). The cohort was slightly male-predominant (51.8%). Hindustani patients comprised the largest ethnic group (52.8%), followed by Creole (19.4%), Other (12.7%), Javanese (10.3%), and Maroon patients (4.9%).

**Table 1:** Baseline Characteristics.

| Variable | Category | n | % |
|---|---|---|---|
| Total cohort |  | 84,691 | 100 |
| Diagnosis | Stroke | 24,584 | 29.0 |
|  | HF | 17,806 | 21.0 |
|  | IHD | 42,301 | 50.0 |
| Sex | Male | 43,897 | 51.8 |
|  | Female | 40,794 | 48.2 |
| Ethnicity | Hindustani | 44,695 | 52.8 |
|  | Creole | 16,424 | 19.4 |
|  | Other | 10,733 | 12.7 |
|  | Javanese | 8,690 | 10.3 |
|  | Maroon | 4,149 | 4.9 |

The annual number of admissions increased markedly over the study period, more than doubling between 2013 and 2022. This finding indicates a substantial increase in the volume of hospitalizations for these major cardiovascular conditions over time, although changes in population structure, healthcare utilization, referral patterns, and access to hospital care may also have contributed to the observed increase.

Patients admitted with Stroke and HF were older on average than those admitted with IHD, while length of stay was longest among patients with Stroke. In absolute terms, the highest number of in-hospital deaths occurred among patients admitted with Stroke, followed by HF and IHD.

### Hospital Admission Volume and Capacity Implications

The volume of hospital admissions for cardiovascular conditions increased markedly over the study period. Annual admissions to the three participating hospitals increased from approximately 5,600 in 2013 to 12,400 in 2022, representing a 121% increase over 10 years. This escalation occurred alongside modest population growth (approximately 1.4% annually) and suggests that the increase in admissions reflects both demographic change and rising age-specific disease incidence(15). Projected forward, if historical admission trends continue unchanged, the three participating hospitals would need to accommodate approximately 24,800 annual cardiovascular admissions by 2030—more than double the baseline. This scale of demand would place substantial pressure on emergency department, cardiology, neurology, and intensive care capacities. The concurrent improvement in in-hospital mortality (Table 2) demonstrates that the health system has managed quality-of-care improvements despite rising volume. However, the combination of rising demand and fixed infrastructure suggests that this dual improvement (more patients + lower mortality) may not be sustainable indefinitely without investment in capacity expansion. These trends underscore the urgency of primary prevention. The HEARTS Initiative’s goal of reducing hypertension prevalence and improving hypertension control represents a direct pathway to reducing the future incidence of acute cardiovascular events, thereby slowing the rate of demand escalation on inpatient services.

**Table 2:** Crude case fatality rates by Ethnicity.

| Ethnic Group<sup>1</sup> | Stroke (%) | HF (%) | IHD (%) |
|---|---|---|---|
| Hindustani | 5.84 | 6.84 | 10.00 |
| Maroon | 6.95 | 8.31 | 8.02 |
| Creole | 7.51 | 7.55 | 11.30 |
| Javanese | 9.78 | 9.32 | 9.60 |
| Other | 7.83 | 6.86 | 6.99 |
| **Total** | **7.09** | **7.33** | **9.74** |

### Crude case fatality rates

Crude case fatality rates (CFRs) differed across disease categories and ethnic groups (Table 2). Overall, the CFR was highest among patients admitted with IHD (9.74%), followed by HF (7.33%) and Stroke (7.09%). Substantial ethnic variation was observed within each disease category (*Table 1*). Among patients with Stroke, the highest CFR was observed among Javanese patients (9.78%), compared with 5.84% among Hindustani patients. A similar pattern was observed for HF, for which Javanese patients had the highest CFR (9.32%), whereas the CFR was lowest among Hindustani patients (6.84%). For IHD, Creole patients had the highest CFR (11.30%), while patients classified as Other had the lowest CFR (6.99%). Hindustani patients consistently had relatively low CFRs across all three disease categories, with rates of 5.84% for Stroke, 6.84% for HF, and 10.00% for IHD.

These crude findings demonstrate that both the overall risk of in-hospital mortality and the magnitude of ethnic differences varied by cardiovascular condition, providing a rationale for examining diagnosis-specific temporal trends and adjusted associations in the multivariable analyses.

### System-Level Progress

Between 2013 and 2022, in-hospital mortality for acute cardiovascular conditions declined significantly, with disease-specific improvement patterns (Figure 1). HF: 14% annual mortality reduction (aOR 0.858, 95% CI 0.841–0.875), IHD: 6% annual reduction (aOR 0.941, 95% CI 0.914–0.969), Stroke: 5% annual reduction (aOR 0.947, 95% CI 0.931–0.963). These improvements suggest system-level advances in acute cardiovascular recognition, referral, or management. The steeper decline in heart failure may reflect expanded access to inpatient care, improved pharmacotherapy, or better recognition of acute decompensation.

**Figure 1:**
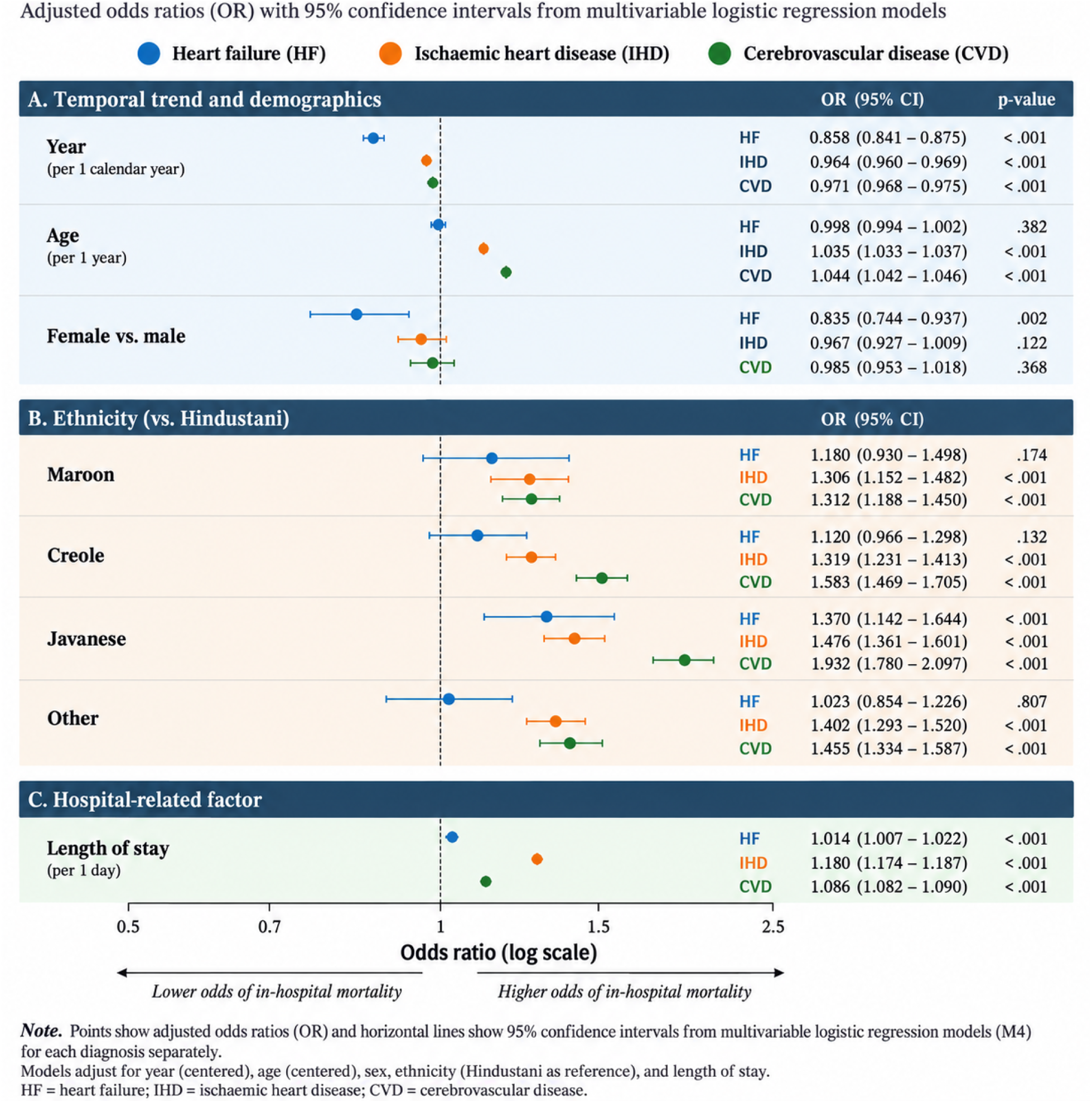
Predictors of in-hospital mortality for HF, IHD and Stroke

### Persistent Equity Gaps

Despite overall system improvement, in-hospital mortality gains were *not* evenly distributed. Ethnicity emerged as a strong predictor of outcome—but in a disease-specific pattern that reveals distinct system gaps:

#### Stroke

Javanese patients experienced substantially higher mortality (aOR 1.828, 95% CI 1.571–2.128) compared with Hindustani patients, as did Creole (aOR 1.393) and other ethnic groups. This ethnic gradient was **most pronounced for stroke**, suggesting inequities in: (a) time to hospital arrival, (b) access to acute stroke care, or (c) quality of in-hospital stroke management.

#### IHD

Ethnic disparities in IHD mortality were smaller and less consistent than for stroke, with elevated mortality in Javanese (aOR 1.558) but not other groups.

#### HF

Only Javanese patients showed elevated mortality (aOR 1.370); other ethnic groups had similar outcomes to Hindustani patients.

### Age and Sex Patterns

Increasing age was strongly associated with mortality in IHD (aOR 1.040 per year) and stroke (aOR 1.018), but not HF. This suggests that older patients with acute coronary syndromes or stroke face particular risk—possibly reflecting delayed presentation, greater comorbidity, or less aggressive acute management in older adults. Female sex was associated with *lower* mortality in HF (aOR 0.835), but no sex differences were observed for IHD or stroke. This may reflect sex-specific patterns in disease severity at presentation or treatment access.

### Discussion Principal findings

This study examined ten years of hospital-based cardiovascular mortality data from three major hospitals in Suriname and identified three principal findings. First, in-hospital mortality declined significantly between 2013 and 2022 for HF, IHD, and Stroke, although the magnitude of decline differed substantially between conditions. Second, age and sex were associated with mortality in disease-specific patterns: age was associated with higher mortality in IHD and Stroke, whereas female sex was associated with lower mortality in HF. Third, ethnic inequalities were particularly pronounced for Stroke. Compared with Hindustani patients, all other major ethnic categories had significantly higher adjusted odds of in-hospital mortality. Taken together, these findings demonstrate that overall cardiovascular mortality trends do not represent a homogeneous phenomenon. Improvements occurred across conditions, but the distribution of mortality differed according to disease and population group.

### Declining mortality across cardiovascular conditions

The decline in in-hospital mortality across all three conditions represents an important temporal finding. However, the observational nature of the study means that the reduction cannot be attributed to a particular treatment, policy, or health-system intervention. Several mechanisms may plausibly contribute to changes in hospital mortality over time, including changes in clinical management, access to care, referral patterns, diagnostic practices, patient characteristics, admission thresholds, and coding practices(16). Because detailed clinical data were unavailable, these mechanisms cannot be distinguished in the present analysis. The different magnitude of decline across conditions is nevertheless informative. HF showed the steepest decline, whereas IHD and Stroke showed more modest reductions(17). This heterogeneity suggests that cardiovascular mortality should be monitored at the disease level rather than through a single aggregate indicator. The finding is particularly relevant for public-health surveillance because an overall improvement could conceal areas in which progress is slower.

### Increasing hospital volume alongside declining mortality

The increase in annual admissions is another important feature of the observed period. Admissions to the participating hospitals more than doubled between 2013 and 2022. This finding should not be interpreted as evidence of increasing disease incidence because hospital admissions are influenced by multiple factors beyond disease occurrence. Population ageing, changes in healthcare-seeking behavior, referral patterns, accessibility, and admission practices may all contribute(18).Nevertheless, the combination of increasing hospital volume and declining in-hospital mortality indicates that the burden placed on hospitals increased substantially while short-term outcomes improved. From a health-system planning perspective, this emphasizes the importance of considering both volume and outcomes. Monitoring mortality alone may underestimate pressure on hospital services, whereas monitoring admissions alone does not capture whether outcomes are improving. The increasing volume also strengthens the rationale for prevention and early management of cardiovascular risk factors(16–19). However, the present study cannot quantify the contribution of prevention to the observed admission trends.

### Age-related inequalities

The strong association between increasing age and mortality among patients hospitalized with IHD is consistent with the vulnerability of older patients to adverse cardiovascular outcomes(18–20).

A similar, although smaller, association was observed for Stroke. These findings may reflect greater comorbidity, more severe disease, physiological vulnerability, or differences in presentation and treatment among older patients(2,21). However, the available data do not allow these mechanisms to be evaluated.

The absence of an independent association between age and HF mortality is noteworthy. HF is clinically heterogeneous, and the administrative data did not permit differentiation by phenotype, severity, or etiology(22). The lack of association should therefore not be interpreted as evidence that age is clinically unimportant in the presence of other risk factors in HF. From a public-health perspective, the age gradients observed for IHD and Stroke reinforce the importance of prevention, risk-factor management, and appropriate access to care among older adults.

### Sex differences were condition-specific

Female sex was associated with lower odds of in-hospital mortality among patients hospitalized with HF, while no statistically significant sex differences were observed for IHD or Stroke. The finding does not imply that sex is unimportant for cardiovascular outcomes in IHD or Stroke. Rather, it indicates that after adjustment for the variables available in this dataset, sex was not independently associated with in-hospital mortality in those two disease categories(23). Potential explanations for sex-specific differences may include differences in age, disease phenotype, comorbidity, presentation, healthcare utilization, or treatment (22–25). These factors could not be adequately assessed in the current administrative dataset. The results therefore support continued sex-disaggregated cardiovascular surveillance rather than assuming that temporal improvements or mortality risks are equivalent between men and women.

### Ethnic inequities in cardiovascular mortality

The most important health-equity finding was the pronounced ethnic gradient in Stroke mortality. Compared with Hindustani patients, mortality was significantly higher among Maroon, Creole, Javanese, and Other ethnic groups. The association was strongest among Javanese patients, whose adjusted odds of in-hospital mortality were approximately 1.8 times those of Hindustani patients. Importantly, ethnicity should not be interpreted as a biological cause of these differences. Ethnic categories are social and population classifications that may capture differences in socioeconomic circumstances, geographic location, healthcare access, health-seeking behavior, risk-factor exposure, continuity of care, or other structural determinants(26,27). The present study cannot determine which of these mechanisms explains the observed disparities. The appropriate interpretation is therefore that ethnicity identifies a population-level inequity signal that warrants further investigation.

### Why the ethnic gradient may be particularly pronounced for cerebrovascular disease

The stronger ethnic gradient observed for Stroke than for HF or IHD is potentially important. Cerebrovascular outcomes can be influenced by processes occurring before hospital admission, including recognition of neurological symptoms, decision-making about seeking care, transportation, geographic accessibility, and time to hospital presentation. Once patients reach hospital, outcomes may also depend on diagnostic capacity, early neurological assessment, access to imaging, specialized expertise, and subsequent management. These factors provide plausible pathways through which structural and healthcare inequalities could contribute to differences in mortality (28). However, none of these mechanisms was measured directly in the present study. The findings therefore should not be interpreted as demonstrating that ethnic differences in Stroke mortality are caused by delayed presentation, unequal access to stroke services, or differences in quality of care. Instead, the observed pattern identifies a priority for future research. This distinction is particularly important because the ethnic gradient was broader for Stroke than for HF. The consistency of the association across four non-reference ethnic groups suggests that the finding is unlikely to be explained solely by a difference between one specific ethnic group and the reference population.

### Implications for public health and health-system planning

The findings have several implications for cardiovascular health policy in Suriname. First, cardiovascular surveillance should routinely report outcomes by disease, age, sex, and ethnicity. Aggregate mortality measures alone may conceal important inequalities. Second, the ethnic disparities observed for Stroke should be investigated using data capable of examining the pathways between population characteristics and outcomes. These should include, where feasible, time from symptom onset to presentation, geographic accessibility, emergency transport, disease severity, diagnostic investigations, treatment, and hospital-level characteristics. Third, the disease-specific differences in temporal trends suggest that cardiovascular policy should not rely exclusively on aggregate cardiovascular indicators. HF, IHD, and Stroke may require different approaches to prevention, early recognition, referral, and care. Fourth, the substantial increase in hospital admission volume highlights the importance of strengthening prevention and risk-factor control. Hypertension is particularly relevant because it is a major modifiable risk factor for both ischemic and cerebrovascular disease. Finally, the findings provide a useful pre-implementation reference point for Suriname’s cardiovascular prevention efforts, including the HEARTS initiative. HEARTS implementation began after the study period; therefore, the present study cannot evaluate its effectiveness. Future monitoring can nevertheless assess whether improvements in cardiovascular outcomes continue and whether those improvements are equitably distributed across population groups.

### Strengths

This study has several strengths. First, it included 84,691 hospital admissions over a ten-year period, providing a substantial longitudinal dataset in a setting where population-based cardiovascular surveillance is limited. Second, the participating hospitals represented approximately 71.8% of national hospital admissions, providing broad coverage of hospital-based cardiovascular care in Suriname. Third, the analysis examined HF, IHD, and Stroke separately. This allowed identification of disease-specific temporal and demographic patterns that would have been obscured by combining heterogeneous cardiovascular conditions. Fourth, ethnicity was explicitly incorporated into the analysis, allowing identification of population-level differences that would not have been apparent from overall mortality estimates. Fifth, multivariable logistic regression allowed adjustment for age, sex, and ethnicity when examining temporal trends and mortality differences.

### Limitations

Several limitations should be considered. First, this was a hospital-based study. Although the participating hospitals account for a large proportion of national admissions, the findings cannot be interpreted as estimates of population-level cardiovascular mortality or incidence. Second, the unit of analysis was the hospital admission rather than the individual patient. Patients with multiple admissions could therefore contribute more than one observation. If repeated admissions from the same individuals were present, conventional logistic regression may underestimate uncertainty. Third, diagnoses were based on routinely collected ICD-10 administrative data. Misclassification is possible, and changes in coding practices over time could influence observed temporal trends. Fourth, the dataset lacked several important clinical variables, including disease severity, comorbidities, vital signs, laboratory results, imaging findings, treatment, and time from symptom onset to hospital presentation. Consequently, the models had limited explanatory power and residual confounding is likely. Fifth, in-hospital mortality captures deaths occurring during hospitalization only. The study therefore cannot assess post-discharge mortality, long-term survival, or total population mortality. Sixth, ethnicity was obtained from routinely collected administrative categories. Misclassification is possible, and broad categories may conceal substantial heterogeneity within population groups. Seventh, the observational design prevents causal inference. In particular, the observed decline in mortality over time cannot be attributed to specific changes in clinical care, health policy, prevention programs, or other interventions. Finally, LOS was not used as a primary explanatory variable because it is determined after admission and can be affected by the outcome itself. Associations between LOS and mortality should therefore not be interpreted as causal effects.

### Future research

The findings provide several directions for further research. First, future studies should examine whether ethnic differences in Stroke mortality are associated with differences in risk-factor control, disease severity, time to presentation, geographic accessibility, emergency transport, diagnostic assessment, or treatment. Second, linkage of hospital records with primary care, emergency medical services, mortality registries, geographic information, and socioeconomic data could provide a more complete assessment of the pathways underlying cardiovascular inequalities. Third, future analyses should examine whether the observed temporal decline in mortality was similar across ethnic and sex groups. Such analyses would directly address whether improvements in cardiovascular outcomes have been equitable. Fourth, incorporation of clinical variables into hospital databases would permit more robust risk adjustment and facilitate monitoring of risk-adjusted mortality over time. Finally, the 2013–2022 data provide a useful pre-implementation reference period for subsequent evaluation of cardiovascular prevention and health-system initiatives, including HEARTS. Future studies should assess both overall effects and distributional effects across population groups.

## Conclusion

In-hospital mortality among patients hospitalized with HF, IHD, and Stroke declined significantly in Suriname between 2013 and 2022, but the magnitude and distribution of improvement differed across cardiovascular conditions and population groups. HF showed the steepest temporal decline, whereas IHD and Stroke demonstrated more modest improvements. Age was associated with higher mortality in IHD and Stroke, while female sex was associated with lower mortality in HF. The most prominent inequality was observed for Stroke, for which all major non-reference ethnic groups had higher adjusted odds of in-hospital mortality than Hindustani patients. These differences should not be interpreted as biological effects of ethnicity, but as an important population-level signal of potential social, structural, geographic, or healthcare-related inequalities. The findings demonstrate the importance of monitoring cardiovascular outcomes not only by disease and over time, but also across population groups. Strengthening routine surveillance and investigating the mechanisms underlying ethnic differences should be priorities for cardiovascular public health in Suriname. The 2013–2022 findings also provide a relevant pre-implementation baseline against which future cardiovascular prevention and health-system initiatives can be evaluated.

## Data availability

The data supporting the findings of this study are not publicly available because of restrictions imposed by the Ministry of Health of Suriname. Data may be made available upon reasonable request and with permission from the Ministry of Health.

## Ethics approval and consent to participate

Ethical oversight was provided by the national review body under approval letter CMWO-14, dated March 25, 2026. The study used retrospectively collected, anonymized hospital data. The requirement for informed consent was waived in accordance with applicable national legislation and institutional policy.

## Consent for publication

Not applicable.

## Declaration of artificial intelligence (AI) tool use

Artificial intelligence tools were used create figures and to assist with language editing, including improving wording, sentence structure, and clarity. All authors reviewed and approved the final manuscript and take full responsibility for its content.

## Author contributions

RD conceived the study, interpreted the data, created figures, and drafted the manuscript. JT and RB provided overall scientific guidance and supervision as promotors. RB contributed to manuscript editing. RGS provided institutional support, feedback, and permission to conduct the study. AM and VJ provided and supported data collection. All authors critically reviewed the manuscript, approved the final version, and agree to be accountable for all aspects of the work.

## Footnotes

1 = Ethnicity categories were based on self-reported population groups routinely registered in hospital administrative records.

